# Intensive care unit and hospital mortality for non-COVID critically ill patients before, and during the COVID-19 pandemic in Alberta hospitals: retrospective, observational cohort study

**DOI:** 10.1101/2025.09.25.25336571

**Authors:** Vincent I Lau, Fadi Hammal, Wendy I Sligl, Constantine J Karvellas, Demetrios J. Kutsogiannis, Fernando G Zampieri, M. Elizabeth Wilcox, Kirsten M. Fiest, Daniel J. Niven, Henry T. Stelfox, Danny J. Zuege, Ken Parhar, Sean M. Bagshaw, Oleksa Rewa

**Affiliations:** Department of Critical Care Medicine, Faculty of Medicine and Dentistry, University of Alberta, and Alberta Health Services, Edmonton, Alberta, Canada; Department of Anesthesia and Pain Medicine, Faculty of Medicine and Dentistry, University of Alberta, and Alberta Health Services, Edmonton, Alberta, Canada; School of Public Health, University of Alberta, Edmonton, Alberta, Canada; Department of Critical Care Medicine, Cumming School of Medicine, University of Calgary, and Alberta Health Services, Calgary, Alberta, Canada; Neuroscience and Mental Health Institute, Edmonton, Alberta, Canada

**Author notes:** <u>Corresponding Author:</u> Vincent Lau, Department of Critical Care Medicine, Faculty of Medicine and Dentistry, University of Alberta and Alberta Health Services, 8440 112 Street, Edmonton, Alberta, Canada.

**Keywords:** mortality, non-COVID illness, COVID-19 pandemic, outcomes, non-communicable diseases

## Abstract

**Objectives:** The Coronavirus Disease-2019 (COVID-19) pandemic had indirect impacts on healthcare provided to critically ill intensive care unit (ICU) non-COVID patients and their outcomes. Therefore, we examined the effect of the COVID-19 pandemic on patients with non-COVID critical illness during the pandemic and pre-pandemic epochs.

**Design:** Retrospective, population-based, observational cohort study.

**Setting:** All adult patients admitted to general ICUs in Alberta, Canada, from March 2017-2020 (pre-pandemic period) and March 2020-2023 (pandemic period).

**Data sources:** Data was captured from an integrated critical care clinical information system (eCritical Alberta) and Alberta Health Services (AHS) administrative databases were utilized.

**Measurements and main results:** A total of 80,540 non-COVID patients were admitted to ICUs in Alberta in the period between March 2017 and March 2023, equally distributed between the pandemic (40,196, 50.1%) and pre-pandemic (40,344, 49.9%) periods. For pandemic versus pre-pandemic cohorts, patient mean age was 57.7 ± 16.2 vs 58.5 ± 16.6 years and mean APACHE II score was 19.5 ± 8.8 vs 18.8 ± 8.5, respectively. ICU mortality was higher during the pandemic compared with pre-pandemic (14.2% vs. 11.2%, mean difference [MD]: +3.1%, adjusted odds ratio [OR]: 1.12; 95% CI: 1.05-1.20, p<0.001) after logistic regression adjustment. The hospital mortality was significantly higher in the pandemic vs. the pre-pandemic period (23.6% vs. 15.9%; MD: +7.7%, adjusted OR: 1.91, 95% CI: 1.81-2.02, p<0.001). There was greater ICU (5.95 ± 9.4 vs. 5.35 ± 8.5 days, MD: +0.63 days, 95% CI: 0.47 to 0.72 days, p<0.001), and lower hospital (18.0 ± 28.0 vs. 19.9 ± 33.3 days, MD: -1.89 days, 95% CI: -1.45 to -2.32 days p<0.001) lengths of stay (LOS) during the pandemic compared with pre-pandemic.

**Conclusion:** During the pandemic period, non-COVID patients had worse outcomes, greater adjusted ICU and hospital mortality, and higher resource utilization with increased ICU lengths of stay, when compared with pre-pandemic periods.

## Background

The coronavirus 2019 (COVID-19) pandemic caused by the severe acute respiratory syndrome coronavirus 2 (SARS-CoV-2) was a global health emergency as declared by the World Health Organization (WHO) (1). As of May 5, 2024 when the WHO declared the pandemic over, there were over 750 million confirmed cases and over 7 million deaths been reported globally (2). Correspondingly, Canada has experienced over 4.8 million cases (12% cases, 12 thousand cases per 100,000 population) and almost 56 thousand total deaths (0.14% deaths, 140 deaths per 100,000 population). In Alberta, over 644 thousand cases (12.8% cases, 12,800 cases per 100,000 population) and just over 6000 deaths (0.12% deaths, 120 deaths per 100,000 population) (3, 4). There is evidence to suggest that non-COVID-19 patients suffered indirect harm during the COVID-19 pandemic, with increased mortality, morbidity, disruptions in care, and changes to hospitalization patterns during the pandemic, when compared to pre-pandemic epochs (5, 6). One hypothesis for these observations may relate to the “rule of rescue” where the most acute and sickest patients who require intensive care unit (ICU) and hospital resources (COVID-19 pandemic patients) are triaged ahead of non-COVID-19 patients (e.g., scheduled surgeries, non-urgent medical illnesses) (5–7). Patients may have also avoided healthcare settings from fear of COVID contraction, potentially worsening outcomes from delayed presentations for care (e.g. acute coronary syndromes) (8–10).

Overall, following mortality and morbidity attributable to COVID-19 alone does not accurately estimate the full impact of the COVID-19 pandemic on the healthcare system. It is crucial to evaluate total excess (e.g., avoidable or unanticipated) mortality (inclusive of both COVID-19 and non-COVID patients) during the pandemic as compared to non-pandemic times (5, 6, 11, 12).

Therefore, our study aims to describe and compare the demographic, clinical characteristics, and outcomes (ICU and hospital mortality) of the non-COVID adult population admitted to ICUs in Alberta during the pandemic (March 2020-2023) and pre-pandemic periods (March 2017-2020).

## Methods

We followed the STrengthening the Reporting of OBservational studies in Epidemiology (STROBE) for reporting observational studies (13). Ethics approval was obtained from the University of Alberta Health Research Ethics Board (File # PRO00130042, approved April 14, 2023).

### Design, setting, and population

This was a retrospective, population-based, cohort study. We included all adult patients admitted to any of the general mixed medical/surgical and subspecialized adult ICUs or all adult patients in pediatric ICUs, who were cared for by adult intensivists in Alberta, Canada from March 5, 2020 (date of first COVID-19 ICU admission in Alberta) to March 5, 2023 (3 years since start of pandemic in Alberta). Our control period (prior to the pandemic) includes all adult patients admitted to the same ICUs from March 5, 2017, to March 4, 2020.

### Data collection and sources

Data collected included demographics (e.g., age, sex), primary diagnosis, admission source, Clinical Frailty Score (CFS) (14), patient illness severity (e.g., Acute Physiology and Chronic Health Evaluation (APACHE) II and III (15), admission Sequential Organ Failure Assessment (SOFA) (16) for adults), organ support therapy (e.g., vasopressors, invasive/non-invasive ventilation, prone positioning, renal replacement therapy, etc.) and in-hospital complications (e.g., nosocomial infections, bleeding, stroke), and patient outcomes (e.g. death in the ICU and in hospital, hospital discharge destination).

Data sources include eCritical Alberta/TRACER and Connect Care/Enterprise/Alberta Health Services and Analytics data repositories (17, 18), which provided demographics, admissions, discharges/transfers, vital statistics and mortality, Discharge abstract database (DAD), diagnostic/case-mix, illness severity: APACHE II and III scores; admission SOFA score), health care utilization in Alberta, including organ supports and lengths of stay (18).

### Outcomes

The primary outcome was mortality (Hospital and ICU). Secondary outcomes included 1) ICU and hospital lengths of stay, 2) hospital discharge destination, and 3) healthcare resource utilization.

### Reporting and Analysis

Descriptive statistics were tabulated according to the relation to COVID-19 (pre-COVID and during COVID-19). Continuous data were reported as means with standard deviations (SD) and compared using t-tests. Median and interquartile ranges (IQR) were reported for non-normally distributed variables and compared using the Wilcoxon rank-sum test. Analysis was performed on complete cases only (those with data included). Categorical variables were presented as counts or proportions and compared using the Chi-square test.

Significance was set to p<0.05. Variables were assessed using clinical rationale to be included in a multinomial regression analysis (19) to estimate the adjusted OR (aOR) for ICU and hospital mortality. Candidate covariables were age (categorized), sex, admission APACHE II, admission class, histories of respiratory insufficiency, diabetes mellitus, liver disease, and heart disease.

Subgroup analysis was performed for age groups (10-year tranches, based on prior reporting of age subgroups in COVID-19 (20) and sex; SOFA score, APACHE II score, APACHE III score, and CFS score for patients. For subgroup analysis, the score cut points were selected based on cut points used in previous studies. For APACHE II ≥ 17 (21), APACHE III ≥ 55 (22), SOFA score ≥ 5 (23), and Clinical Frailty Scale ≥ 4 (24).

We also performed propensity-score matching (PSM) (25) cohort comparisons using a logistic regression model that included the same set of covariables used in the logistic regression analysis (listed above). Patients in the COVID period were matched 1:1 to those in pre-COVID, using nearest neighbor matching without replacement (25). Following PSM, baseline demographics, clinical characteristics and outcomes were re-evaluated using regression models for the matched cohort. For comparison, we also evaluated and described the remaining unmatched cohort as well.

## Results

### Baseline demographics and clinical characteristics

Overall, there were 80,540 non-COVID patients admitted to ICUs in Alberta in the period between March 2017 and March 2023, relatively equally distributed between pre-COVID (n=40,196, 50.1%) and during the COVID-19 pandemic period (n=40,344, 49.9%), not including expanded capacity in the dedicated COVID-19 ICUs (as non-COVID patients were not admitted there0. Table 1 shows the characteristics of patients admitted to ICU. Age and sex distribution were similar. There was no significant difference between pre-pandemic and pandemic periods in mean admission SOFA score [6.5 ± 3.9 vs 6.4 ± 3.9, p=0.51]. However, there were significant differences in mean weight [84.5 ± 24.5 vs. 86.2 ± 25.7 kg, p<0.001], APACHE II score [84.5 ± 24.5 vs. 86.2 ± 25.7, p<0.001], APACHE III score [p<0.001], and CFS score [p<0.001] in pre-pandemic vs. pandemic periods. (Table 1). To adjust for potential confounding, we also performed propensity-matching using a logistic multivariable regression model based on age and illness severity (APACHE II, SOFA, CFS). We present both the propensity-matched (Supplemental Table 1) and non-propensity matched cohorts (Supplemental Table 2).

### Admission geographic location and subtypes

Supplemental Table 3 shows the distribution of ICU admissions by geographic hospital location, specialty (e.g., general, neurologic vs. cardiovascular), and academic vs. community subtypes. Subspecialized ICUs (e.g., cardiovascular and neurological) saw decreased admissions during the pandemic compared to pre-pandemic periods. There was no distinct trend of increased or reduced utilization between academic versus community hospitals.

### Admission diagnosis subtypes

Supplemental Table 4 shows the types of ICU admissions during the pandemic versus the pre-pandemic periods. Of note, there were significantly fewer elective operations during the pandemic compared to pre-pandemic periods. There was a significant increase in medical and ICU admissions of various subtypes (e.g., cardiovascular, gastrointestinal, hematology, genitourinary, metabolic, musculoskeletal, neurological, pulmonary, transplant, and trauma).

### Mortality

The ICU mortality rate was significantly higher in the pandemic period vs. the pre-pandemic period for non-COVID ICU patients (Figure 1). Crude ICU mortality odds ratio was 14.2% vs. 11.2% (MD: +3.0%, OR: 1.31 (95% CI: 1.26 to 1.37), p<0.001), and adjusted mortality (aOR) was: 1.21 (95% CI: 1.14 to 1.28, p<0.001) when adjusted for age, sex, weight, APACHE II score, admission class, respiratory insufficiency, diabetes, liver disease and heart disease. . Similarly, the hospital mortality rate was significantly higher in pandemic period vs. the pre-pandemic period (Figure 1), with 23.6% vs. 15.9% (MD: +7.7%, OR: 1.63, 95% CI: 1.58 to 1.69, p<0.001) for crude mortality rate, and adjusted mortality (aOR): 2.06, 95% CI: 1.97 to 2.16, p<0.001) (Table 2).

### Length of stay

The length of stay (LOS) in ICU was significantly higher in the pandemic period compared to the pre-pandemic period for non-COVID patients (3.05 [1.3, 6.8] vs 2.96 [1.4, 6.0], <0.001) . In contrast, the hospital was significantly lower in the pandemic period vs. the pre-pandemic period (9.0 [5.0, 20] vs 10 [5.0, 21], <0.001) (Table 2).

### Subgroup mortality analysis

Table 3 demonstrates various subgroup differences in mortality between the pre-pandemic and pandemic non-COVID critically ill patient cohorts, alongside subgroup forest plot in Figures 1a and 1b. ICU and hospital mortality rates were significantly higher during the pandemic compared to the pre-pandemic periods for the majority of subgroups: age 18-90, any sex, any SOFA, APACHE and CFS score.

### Resource Use and Organ Supports

Supplemental Table 5 shows the resource utilization, including ICU level organs supports (e.g. ventilation, vasopressors, dialysis). There was a significant increase in the proportion of non-COVID ICU patients who needed invasive ventilation [71.7% vs 66.8%, p<0.001] and median days of invasive ventilation in the pandemic period vs. pre-pandemic period (median: 1.6 days [IQR: 0.5-5.1] vs. 1.0 days [0.3-3.5], p<0.001. There was also significant difference in days of utilization on an average per-patient basis for non-invasive ventilation, continuous renal replacement, but no difference in the hours of intermittent hemodialysis.

## Discussion

In this retrospective, observational cohort study, we found an increase in crude and adjusted ICU and hospital mortality for patients with non-COVID during the COVID-19 pandemic compared to historical non-pandemic epochs. This observation was consistent across all subgroups, including age, sex, and illness severity scores. There was also longer ICU LOS but shorter hospital LOS during the pandemic vs. the pre-pandemic period, likely secondary to increased mortality on the hospital wards.

This analysis provides insight into the potential trade-offs that have occurred for both patients with non-COVID illness and health systems’ capacity to meet standards of care. In multiple jurisdictions, excess all-cause mortality (USA: 72 deaths per 100,000 patient population, UK: 95 deaths/100,000, Spain: 102 deaths/100,000) has been reported over and above recorded COVID-19 deaths alone (26). Multiple jurisdictions have reported increased ICU and hospital mortality for non-COVID critically ill patients as compared to historical norms (5, 6, 27–36). There is emerging literature that excess mortality is not only driven by COVID-19 deaths (6), but there is also non-COVID excess mortality and morbidity (5), including in intensive care units (36), potentially secondary to disruptions of global healthcare services by the COVID-19 pandemic (5, 6). The intensity of disruption (severity multiplied by duration) may have altered the apparent effects among non-COVID illnesses, leading to the variability observed for different jurisdictions and illnesses. This highlights that the pandemic itself has had unintended damage to the normal functioning of our health systems and negatively impacted outcomes for non-COVID patients (5, 6).

Due to the “rule of rescue”, which states that we use available resources to save the life in front of us, even if those resources might be better used elsewhere (7, 37). This can be at the expense of other patients. Because COVID-19 patients often had the highest acuity of illness, resources may have been preferentially directed to them, potentially at the expense of non-COVID critically ill patients. In healthcare, this tension arises between the injunction to do as much good as possible with scarce resources and the injunction to rescue identifiable individuals in immediate peril, regardless of cost. This tension can generate serious ethical and political difficulties for healthcare public policy makers faced with making explicit decisions about public funding (38).

There may be ongoing challenges. There has been evidence of increased ICU healthcare worker turnover (39–43), which can further compromise patient care going forward post-pandemic. There is evidence demonstrating increased association with mortality when ICU staffing to patient ratios is compromised (44–54) where patient-to-nurse ratios >2:1 are associated with increased adjusted relative risk (RR) of mortality of 2.3, and ratios >2.5:1 are associated with adjusted RR of 3.5 (55). Given this association, it will be important to monitor ICU healthcare worker turnover and to see if ICU mortality reduces back to historical norms or if they remain elevated post-pandemic.

The strengths of our study include adherence to STROBE guidelines (13), presenting both crude and adjusted mortality for age, and using a multiple illness severity score to ensure proper interpretation of baseline cohort differences using logistic regression analysis. We also have extended the reference pre-pandemic cohort and pandemic cohorts to three years to observe both the pandemic and post-pandemic trends in mortality, as well as provide a more robust pre-pandemic control cohort to ensure proper adjustment of baseline prognostic risk factors. The data repositories covering all Alberta ICUs in the province (population-based study) are also a strength of this study. We are also one of the only studies to report on sedative, analgesic, and paralytic medication use during the pandemic period on non-COVID patients, which may explain the increased ventilator days for this cohort compared to pre-pandemic periods.

This study also has several limitations. First, we do not have data on morbidity or health-related quality of life in critically ill ICU patients from the registry population-level dataset, as mortality is not the only patient-important outcome to be studied. Second, we do not currently have capacity strain, bed census and occupancy or healthcare provider (e.g. registered nurses) to patient ratios data to supplement our analysis as a marker of potential reasons for increased mortality; however, this is an area of future research to determine if there are correlations between these capacity strain, ICU staffing, and increased mortality. Third, despite Alberta having a robust, single provincial repository for data, there was still a large proportion of patients who had missing data, potentially as a product of how busy clinicians were during the COVID-19 pandemic, that routine data capture and charting were foregone because of heightened patient care duties. Fourth, despite our adjustment when we looked at mortality, there are likely other confounders that were not accounted for during logistic regression modeling analysis, which may impact the results.

## Conclusion

The COVID-19 pandemic had effects on non-COVID illness, including increased ICU and hospital mortality (both crude and adjusted), increased ICU length of stay, but decreased hospital length of stay (perhaps secondary to increased death) in Alberta ICUs and hospitals. Informing public healthcare policy and decision-makers of the potential pandemic effects is crucial to mitigate the impact of the COVID-19 pandemic on both COVID-19 and non-COVID patients, in particular, excess mortality, morbidity, and disruptions in care.

## Data Availability

All data produced in the present work are contained in the manuscript

## Notes

### Competing Interest Statement

The authors have declared no competing interest.

### Funding Statement

This study did not receive any funding

### Author Declarations

Ethics committee of University of Alberta gave ethical approval for this work (Pro00114411, approved: September 27, 2021)

## References

1. Ciotti M, Ciccozzi,Massimo, Terrinoni,Alessandro, et al.: The COVID-19 pandemic. Critical Reviews in Clinical Laboratory Sciences 2020; 57:365–388

2. Sarker R, Roknuzzaman ASM, Nazmunnahar, et al.: The WHO has declared the end of pandemic phase of COVID-19: Way to come back in the normal life. Health Sci Rep 2023; 6:e1544

3. Canada PHA of: COVID-19 daily epidemiology update: Key updates [Internet]. Aem 2020; [cited 2025 Jun 24] Available from: https://www.canada.ca/en.html

4. Alberta COVID-19 Opioid Response Surveillance Report Q2 2020. 2020;

5. Lau VI, Dhanoa S, Cheema H, et al.: Non-COVID outcomes associated with the coronavirus disease-2019 (COVID-19) pandemic effects study (COPES): A systematic review and meta-analysis. PLOS ONE 2022; 17:e0269871

6. Lu D, Dhanoa S, Cheema H, et al.: Coronavirus disease 2019 (COVID-19) excess mortality outcomes associated with pandemic effects study (COPES): A systematic review and meta-analysis [Internet]. Frontiers in Medicine 2022; 9[cited 2023 May 19] Available from: https://www.frontiersin.org/articles/10.3389/fmed.2022.999225

7. März JW, Holm S, Schlander M: Resource allocation in the Covid-19 health crisis: are Covid-19 preventive measures consistent with the Rule of Rescue? Med Health Care Philos 2021; 24:487–492

8. Kite TA, Pallikadavath S, Gale CP, et al.: The Direct and Indirect Effects of COVID-19 on Acute Coronary Syndromes. Heart Fail Clin 2023; 19:185–196

9. Ball S, Banerjee A, Berry C, et al.: Monitoring indirect impact of COVID-19 pandemic on services for cardiovascular diseases in the UK. Heart 2020; 106:1890–1897

10. Ball J, Nehme Z, Bernard S, et al.: Collateral damage: Hidden impact of the COVID-19 pandemic on the out-of-hospital cardiac arrest system-of-care. Resuscitation 2020; 156:157–163

11. Anzai T, Fukui K, Ito T, et al.: Excess mortality from suicide during the early COVID-19 pandemic period in Japan: a time-series modeling before the pandemic. J Epidemiol 2020;

12. Banerjee A, Pasea L, Harris S, et al.: Estimating excess 1-year mortality associated with the COVID-19 pandemic according to underlying conditions and age: a population-based cohort study. The Lancet 2020; 395:1715–1725

13. von Elm E, Altman DG, Egger M, et al.: The Strengthening the Reporting of Observational Studies in Epidemiology (STROBE) statement: guidelines for reporting observational studies. J Clin Epidemiol 2008; 61:344–349

14. Rockwood K, Song X, Mitnitski A: Changes in relative fitness and frailty across the adult lifespan: evidence from the Canadian National Population Health Survey. CMAJ 2011; 183:E487–494

15. Knaus WA, Draper EA, Wagner DP, et al.: APACHE II: A severity of disease classification system. Critical Care Medicine 1985; 13:818–829

16. Vincent JL, Moreno R, Takala J, et al.: The SOFA (Sepsis-related Organ Failure Assessment) score to describe organ dysfunction/failure. On behalf of the Working Group on Sepsis-Related Problems of the European Society of Intensive Care Medicine. Intensive Care Med 1996; 22:707–710

17. Sandhu N, Whittle S, Southern DA, et al.: Health data governance for research use in Alberta. Int J Popul Data Sci 8:2160

18. Bowker SL, Stelfox HT, Bagshaw SM: Critical Care Strategic Clinical Network: Information infrastructure ensures a learning health system. CMAJ 2019; 191:S22–S23

19. Zabor EC, Reddy CA, Tendulkar RD, et al.: Logistic Regression in Clinical Studies. International Journal of Radiation Oncology, Biology, Physics 2022; 112:271–277

20. Signorelli C, Odone A: Age-specific COVID-19 case-fatality rate: no evidence of changes over time. Int J Public Health 2020; 65:1435–1436

21. Tian Y, Yao Y, Zhou J, et al.: Dynamic APACHE II Score to Predict the Outcome of Intensive Care Unit Patients. Front Med (Lausanne) 2022; 8:744907

22. Cho DY, Wang YC: Comparison of the APACHE III, APACHE II and Glasgow Coma Scale in acute head injury for prediction of mortality and functional outcome. Intensive Care Med 1997; 23:77–84

23. Baradari AG, Firouzian A, Davanlou A, et al.: COMPARISON OF PATIENTS’ ADMISSION, MEAN AND HIGHEST SOFA SCORES IN PREDICTION OF ICU MORTALITY: A PROSPECTIVE OBSERVATIONAL STUDY. Mater Sociomed 2016; 28:343–347

24. Bruno RR, Wernly B, Bagshaw SM, et al.: The Clinical Frailty Scale for mortality prediction of old acutely admitted intensive care patients: a meta-analysis of individual patient-level data. Ann Intensive Care 2023; 13:37

25. Ross ME, Kreider AR, Huang Y-S, et al.: Propensity Score Methods for Analyzing Observational Data Like Randomized Experiments: Challenges and Solutions for Rare Outcomes and Exposures. American Journal of Epidemiology 2015; 181:989–995

26. Bilinski A, Emanuel EJ: COVID-19 and Excess All-Cause Mortality in the US and 18 Comparison Countries [Internet]. JAMA 2020; [cited 2020 Oct 19] Available from: https://jamanetwork.com/journals/jama/fullarticle/2771841

27. Wai AK-C, Yip TF, Wong YH, et al.: The Effect of the COVID-19 Pandemic on Non-COVID-19 Deaths: Population-Wide Retrospective Cohort Study. JMIR Public Health Surveill 2024; 10:e41792

28. Zhong X, Ashiru-Oredope D, Pate A, et al.: Clinical and health inequality risk factors for non-COVID-related sepsis during the global COVID-19 pandemic: a national case-control and cohort study [Internet]. eClinicalMedicine 2023; 66 [cited 2024 Mar 6] Available from: https://www.thelancet.com/journals/eclinm/article/PIIS2589-5370(23)00498-4/fulltext

29. McAlister FA, Chu A, Qiu F, et al.: Outcomes Among Patients Hospitalized With Non–COVID-19 Conditions Before and During the COVID-19 Pandemic in Alberta and Ontario, Canada. JAMA Network Open 2023; 6:e2323035

30. Dang A, Thakker R, Li S, et al.: Hospitalizations and Mortality From Non–SARS-CoV-2 Causes Among Medicare Beneficiaries at US Hospitals During the SARS-CoV-2 Pandemic. JAMA Network Open 2022; 5:e221754

31. Bodilsen J, Nielsen PB, Søgaard M, et al.: Hospital admission and mortality rates for non-covid diseases in Denmark during covid-19 pandemic: nationwide population based cohort study. BMJ 2021; 373:n1135

32. Zampieri FG, Bastos LSL, Soares M, et al.: The association of the COVID-19 pandemic and short-term outcomes of non-COVID-19 critically ill patients: an observational cohort study in Brazilian ICUs. Intensive Care Med 2021; 47:1440–1449

33. Payet C, Polazzi S, Rimmelé T, et al.: Mortality Among Noncoronavirus Disease 2019 Critically Ill Patients Attributable to the Pandemic in France. Crit Care Med 2022; 50:138–143

34. McLarty J, Litton E, Beane A, et al.: Non-COVID-19 intensive care admissions during the pandemic: a multinational registry-based study. Thorax 2024; 79:120–127

35. Leafloor CW, Imsirovic H, Qureshi D, et al.: Characteristics and Outcomes of ICU Patients Without COVID-19 Infection—Pandemic Versus Nonpandemic Times: A Population-Based Cohort Study. Crit Care Explor 2023; 5:e0888

36. Wilcox ME, Rowan KM, Harrison DA, et al.: Does Unprecedented ICU Capacity Strain, As Experienced During the COVID-19 Pandemic, Impact Patient Outcome? Crit Care Med 2022; 50:e548–e556

37. Jonsen AR: Bentham in a box: technology assessment and health care allocation. Law Med Health Care 1986; 14:172–174

38. Cookson R, McCabe C, Tsuchiya A: Public healthcare resource allocation and the Rule of Rescue. Journal of Medical Ethics 2008; 34:540–544

39. Arabi YM, Azoulay E, Al-Dorzi HM, et al.: How the COVID-19 pandemic will change the future of critical care. Intensive Care Med 2021; 47:282–291

40. Ke Y-T, Hung C-H: Predictors of Nurses’ Intent To Continue Working at Their Current Hospital. Nursing Economics 2017; 35:259–266

41. Lambden JP, Chamberlin P, Kozlov E, et al.: Association of Perceived Futile or Potentially Inappropriate Care With Burnout and Thoughts of Quitting Among Health-Care Providers. Am J Hosp Palliat Care 2019; 36:200–206

42. Ramírez-Elvira S, Romero-Béjar JL, Suleiman-Martos N, et al.: Prevalence, Risk Factors and Burnout Levels in Intensive Care Unit Nurses: A Systematic Review and Meta-Analysis. Int J Environ Res Public Health 2021; 18:11432

43. Shoorideh FA, Ashktorab T, Yaghmaei F, et al.: Relationship between ICU nurses’ moral distress with burnout and anticipated turnover. Nurs Ethics 2015; 22:64–76

44. Aiken LH, Clarke SP, Sloane DM, et al.: Hospital Nurse Staffing and Patient Mortality, Nurse Burnout, and Job Dissatisfaction. JAMA 2002; 288:1987–1993

45. Aragon Penoyer D: Nurse staffing and patient outcomes in critical care: A concise review. Critical Care Medicine 2010; 38:1521–1528

46. Dall’Ora C, Maruotti A, Griffiths P: Temporary Staffing and Patient Death in Acute Care Hospitals: A Retrospective Longitudinal Study. J Nurs Scholarsh 2020; 52:210–216

47. Daouda OS, Hocine MN, Temime L: Determinants of healthcare worker turnover in intensive care units: A micro-macro multilevel analysis. PLoS One 2021; 16:e0251779

48. Driscoll A, Grant MJ, Carroll D, et al.: The effect of nurse-to-patient ratios on nurse-sensitive patient outcomes in acute specialist units: a systematic review and meta-analysis. Eur J Cardiovasc Nurs 2018; 17:6–22

49. Falk A-C: Nurse staffing levels in critical care: The impact of patient characteristics. Nurs Crit Care 2023; 28:281–287

50. Kiekkas P, Sakellaropoulos GC, Brokalaki H, et al.: Association between nursing workload and mortality of intensive care unit patients. J Nurs Scholarsh 2008; 40:385–390

51. Lasater KB, Aiken LH, Sloane DM, et al.: Chronic hospital nurse understaffing meets COVID-19: an observational study. BMJ Qual Saf 2021; 30:639–647

52. Lee A, Cheung YSL, Joynt GM, et al.: Are high nurse workload/staffing ratios associated with decreased survival in critically ill patients? A cohort study. Ann Intensive Care 2017; 7:46

53. Joy M, Hobbs FDR, McGagh D, et al.: Excess mortality from COVID-19 in an English sentinel network population [Internet]. The Lancet Infectious Diseases 2020; 0[cited 2020 Oct 2] Available from: https://www.thelancet.com/journals/laninf/article/PIIS1473-3099(20)30632-0/abstract

54. Musy SN, Endrich O, Leichtle AB, et al.: The association between nurse staffing and inpatient mortality: A shift-level retrospective longitudinal study. International Journal of Nursing Studies 2021; 120:103950

55. Neuraz A, Guérin C, Payet C, et al.: Patient Mortality Is Associated With Staff Resources and Workload in the ICU: A Multicenter Observational Study*. Critical Care Medicine 2015; 43:1587–1594

