## Supplementary material for "Intensive care unit and hospital mortality for non-COVID critically ill patients before, and during the COVID-19 pandemic in Alberta hospitals: retrospective, observational cohort study": Figures and Tables

Characteristics of the pre-pandemic period adult patients and non-COVID pandemic period patients in Alberta's Intensive Care Units

**Table 1: Patient characteristics**

|  |  | **Pre-pandemic period**  **n=40,196** | | | | | |  | **Non COVID-pandemic period**  **n=40,344** | | | | | |  | | **p value** | |
| --- | --- | --- | --- | --- | --- | --- | --- | --- | --- | --- | --- | --- | --- | --- | --- | --- | --- | --- |
| **Sex** |  |  | | |  | | |  |  | | |  | | | |  | | **0.011** |
| Female |  | 15537 | | | 38.7% | | |  | 15184 | | | 37.6% | | | |  | |  |
| Male |  | 24653 | | | 61.3% | | |  | 25155 | | | 62.4% | | | |  | |  |
| Missing |  | 6 | | | 0.0% | | |  | 5 | | | 0.0% | | | |  | |  |
| **Age** |  |  | | |  | | |  |  | | |  | | | |  | | **<0.001** |
| 18-29 |  | 2952 | | | 7.3% | | |  | 2788 | | | 6.9% | | | |  | |  |
| 30-39 |  | 3478 | | | 8.7% | | |  | 3888 | | | 9.6% | | | |  | |  |
| 40-49 |  | 4307 | | | 10.7% | | |  | 4789 | | | 11.9% | | | |  | |  |
| 50-59 |  | 7674 | | | 19.1% | | |  | 7714 | | | 19.1% | | | |  | |  |
| 60-69 |  | 10551 | | | 26.2% | | |  | 10878 | | | 27.0% | | | |  | |  |
| 70-79 |  | 8153 | | | 20.3% | | |  | 8025 | | | 19.9% | | | |  | |  |
| 80-89 |  | 2828 | | | 7.0% | | |  | 2118 | | | 5.2% | | | |  | |  |
| 90+ |  | 253 | | | 0.6% | | |  | 144 | | | 0.4% | | | |  | |  |
| **Comorbidities** |  |  | | |  | | |  |  | | |  | | | |  | |  |
| Dialysis |  | 1110 | | | 2.9% | | |  | 948 | | | 2.8% | | | |  | | 0.38 |
| Heart Disease |  | 3602 | | | 9.6% | | |  | 3190 | | | 9.5% | | | |  | | 0.95 |
| Respiratory insufficiency |  | 4236 | | | 11.2% | | |  | 3447 | | | 10.3% | | | |  | | **<0.001** |
| Liver disease |  | 1935 | | | 5.1% | | |  | 2017 | | | 6.0% | | | |  | | **<0.001** |
| Cirrhosis |  | 1702 | | | 4.5% | | |  | 1765 | | | 5.3% | | | |  | | **<0.001** |
| Hepatic failure |  | 909 | | | 2.4% | | |  | 950 | | | 2.8% | | | |  | | **<0.001** |
| Diabetes |  | 8893 | | | 23.6% | | |  | 8181 | | | 24.5% | | | |  | | **0.006** |
| Leukemia/M. Myeloma |  | 302 | | | 0.8% | | |  | 243 | | | 0.7% | | | |  | | 0.28 |
| Lymphoma |  | 312 | | | 0.8% | | |  | 256 | | | 0.8% | | | |  | | 0.36 |
| Immune suppression |  | 2818 | | | 7.5% | | |  | 2607 | | | 7.8% | | | |  | | 0.10 |
| Metastatic cancer |  | 1104 | | | 2.9% | | |  | 845 | | | 2.5% | | | |  | | **<0.001** |
| **Admission class** |  |  | | |  | | |  |  | | |  | | | |  | | **<0.001** |
| Medical |  | 18140 | | | 48.1% | | |  | 18537 | | | 55.5% | | | |  | |  |
| Surgical |  | 15219 | | | 40.4% | | |  | 11396 | | | 34.1% | | | |  | |  |
| Neuro |  | 2713 | | | 7.2% | | |  | 2097 | | | 6.3% | | | |  | |  |
| Trauma and head injury |  | 884 | | | 2.3% | | |  | 713 | | | 2.1% | | | |  | |  |
| Trauma without head injury |  | 725 | | | 1.9% | | |  | 649 | | | 1.9% | | | |  | |  |
| **Clinical Frailty Scale (CFS)** |  |  | | |  | | |  |  | | |  | | | |  | | **<0.001** |
| Very fit |  | 3263 | | | 9.6% | | |  | 2370 | | | 7.5% | | | |  | |  |
| Fit/ Well |  | 5715 | | | 16.7% | | |  | 6144 | | | 19.5% | | | |  | |  |
| Managing Well |  | 7719 | | | 22.6% | | |  | 7543 | | | 23.9% | | | |  | |  |
| Vulnerable |  | 8412 | | | 24.7% | | |  | 8101 | | | 25.7% | | | |  | |  |
| Mildly Frail |  | 3864 | | | 11.3% | | |  | 3335 | | | 10.6% | | | |  | |  |
| Moderately Frail |  | 3256 | | | 9.5% | | |  | 2602 | | | 8.2% | | | |  | |  |
| Severely Frail |  | 1523 | | | 4.5% | | |  | 1137 | | | 3.6% | | | |  | |  |
| Very Severely Frail |  | 265 | | | 0.8% | | |  | 243 | | | 0.8% | | | |  | |  |
| Terminally ill |  | 104 | | | 0.3% | | |  | 88 | | | 0.3% | | | |  | |  |
| **APACHE II score** |  |  | | |  | | |  |  | | |  | | | |  | | <0.001 |
| < 17 |  | 17653 | | | 45.1% | | |  | 16678 | | | 42.2% | | | |  | |  |
| ≥ 17 |  | 21530 | | | 54.9% | | |  | 22833 | | | 57.8% | | | |  | |  |
| **APACHE III score** |  |  | | |  | | |  |  | | |  | | | |  | | 0.63 |
| < 55 |  | 17649 | | | 44.0% | | |  | 17584 | | | 43.8% | | | |  | |  |
| ≥ 55 |  | 22480 | | | 56.0% | | |  | 22553 | | | 56.2% | | | |  | |  |
| **SOFA score** |  |  | | |  | | |  |  | | |  | | | |  | | **0.023** |
| < 5 |  | 13425 | | | 33.4% | | |  | 13758 | | | 34.2% | | | |  | |  |
| ≥ 5 |  | 26758 | | | 66.6% | | |  | 26507 | | | 65.8% | | | |  | |  |
| **CFS score** |  |  | | |  | | |  |  | | |  | | | |  | | **<0.001** |
| < 5 |  | 25109 | | | 73.6% | | |  | 24158 | | | 76.5% | | | |  | |  |
| ≥ 5 |  | 9012 | | | 26.4% | | |  | 7405 | | | 23.5% | | | |  | |  |
|  |  | **n** | **%** | **Mean** | | **(SD)** | **Median [IQR]** |  | **n** | **%** | **Mean** | | **(SD)** | **Median [IQR]** | |  | | **p value**† |
| **Age (year)** |  | 40196 | 100% | 58.49 | | (16.59) | 61 [48,71] |  | 40344 | 100% | 57.65 | | (16.16) | 60 [47, 70] | |  | | **<0.001** |
| **Weight (KG)** |  | 38550 | 95.9% | 84.4 6 | | (24.53) | 81 [68,96.4] |  | 24101 | 59.7% | 86.15 | | (25.68) | 82 [69, 98.4] | |  | | **<0.001** |
| **APACHE-II** |  | 39183 | 97.5% | 18.84 | | (8.45) | 18 [13, 24] |  | 39511 | 97.9% | 19.50 | | (8.82) | 18 [13, 25] | |  | | **<0.001** |
| **Invasive ventilation (day)** |  | 26839 | 66.8% | 3.34 | | (6.99) | 0.99 [0.35-3.5] |  | 28921 | 71.7% | 4.47 | | (7.97) | 1.6 [0.5, 5.1] | |  | | **<0.001** |

APACHE: Acute Physiologic Assessment and Chronic Health Evaluation, CFS: Clinical Frailty Scale, CI: confidence intervals, COVID: Coronavirus Disease, ICU: intensive care unit, KG: kilograms, SD: standard deviation, SOFA: Sequential Organ Failure Assessment, † Mann-Whitney U test

**Table 2: Outcomes**

|  |  | **Pre-pandemic period** | | | |  | **Non COVID-pandemic period** | | | |  |  |  |
| --- | --- | --- | --- | --- | --- | --- | --- | --- | --- | --- | --- | --- | --- |
|  |  | **n/N** | | **%** | |  | **n/N** | | **%** | |  | **OR (95% CI)** | **p value** |
| **ICU mortality** |  |  | |  | |  |  | |  | |  |  |  |
| Crude |  | 4,503/40,196 | | 11.2% | |  | 5,727/40,344 | | 14.2% | |  | 1.31 (1.26, 1.37) | <0.001 |
| Adjusted* |  |  | |  | |  |  | |  | |  | 1.21 (1.14, 1.28) | <0.001 |
| Propensity-matched cohort |  | 2,008/15,614 | | 12.9% | |  | 1,930/15,614 | | 12.4% | |  | 0.96 (0.89, 1.02) | 0.18 |
| Propensity-unmatched cohort |  | 2,495/24,582 | | 10.1% | |  | 3,797/24,730 | | 15.4% | |  | 1.61 (1.52, 1.70) | <0.001 |
| **Hospital mortality** |  |  | |  | |  |  | |  | |  |  |  |
| Crude |  | 6,402/40,196 | | 15.9% | |  | 9,537/40,344 | | 23.6% | |  | 1.63 (1.58, 1.69) | <0.001 |
| Adjusted* |  |  | |  | |  |  | |  | |  | 2.06 (1.97, 2.16) | <0.001 |
| Propensity-matched cohort |  | 2,655/15,614 | | 17.0% | |  | 3,355/15,614 | | 21.5% | |  | 1.34 (1.26, 1.41) | <0.001 |
| Propensity-unmatched cohort |  | 3,747/24,582 | | 15.2% | |  | 6,182/24,730 | | 25.0% | |  | 1.85 (1.77, 1.94) | <0.001 |
|  |  | **Mean (SD)** | | **Median [IQR]** | |  | **Mean (SD)** | | **Median [IQR]** | |  | **Mean Difference**  **(95% CI)** | **p value** |
| **ICU Length of Stay** |  |  |  |  |  |  |  |  |  |  |  |  |  |
| Full cohort |  | 5.35 | (8.51) | 2.96 | [1.4-6.0] |  | 5.95 | (9.44) | 3.05 | [1.3-6.8] |  | 0.63 (0.47, 0.72) | <0.001† |
| Propensity-matched cohort |  | 5.93 | (7.35) | 3.52 | [1.7-7.2] |  | 5.42 | (6.70) | 3.14 | [1.3-6.9] |  | -0.50 (-0.66, -0.35) | <0.001† |
| Propensity-unmatched cohort |  | 4.98 | (9.15) | 2.74 | [1.2-5.2] |  | 6.28 | (10.81) | 3.0 | [1.3-6.7] |  | 1.29 (1.12, 1.47) | <0.001† |
| **Hospital Length of Stay** |  |  |  |  |  |  |  |  |  |  |  |  |  |
| Full cohort |  | 19.88 | (33.26) | 10 | [5-21] |  | 18.00 | (27.99) | 9 | [5-20] |  | -1.89 (-2.32, -1.45) | <0.001† |
| Propensity-matched cohort |  | 20.76 | (33.90) | 10 | [5-22] |  | 16.64 | (24.74) | 9 | [5-18] |  | -4.13 (-4.80, -3.45) | <0.001† |
| Propensity-unmatched cohort |  | 19.32 | (32.84) | 9 | [4-20] |  | 18.85 | (29.82) | 10 | [4-21] |  | -0.48 (-1.04, 0.09) | 0.19† |

CI: confidence intervals, COVID: Coronavirus Disease; ICU: intensive care unit, OR: odds ratio, SD: standard deviation

OR reported for pandemic compared to pre-pandemic

* Adjusted in the full cohort for: Age, sex, weight, APACHE II, admission class, respiratory insufficiency, diabetes, liver disease, heart disease

† Wilcoxon rank-sum test

**Figure 1: Non-COVID ICU and Hospital Mortality**

Figure 1a: Non-COVID ICU Mortality (counts)

ICU Deaths (n)

| Month | ICU Death |
| --- | --- |
| Pre-COVID 1 | 117 |
| Pre-COVID 2 | 114 |
| Pre-COVID 3 | 112 |
| Pre-COVID 4 | 117 |
| Pre-COVID 5 | 128 |
| Pre-COVID 6 | 102 |
| Pre-COVID 7 | 126 |
| Pre-COVID 8 | 117 |
| Pre-COVID 9 | 116 |
| Pre-COVID 10 | 151 |
| Pre-COVID 11 | 160 |
| Pre-COVID 12 | 116 |
| Pre-COVID 13 | 142 |
| Pre-COVID 14 | 113 |
| Pre-COVID 15 | 127 |
| Pre-COVID 16 | 127 |
| Pre-COVID 17 | 99 |
| Pre-COVID 18 | 109 |
| Pre-COVID 19 | 130 |
| Pre-COVID 20 | 123 |
| Pre-COVID 21 | 138 |
| Pre-COVID 22 | 123 |
| Pre-COVID 23 | 123 |
| Pre-COVID 24 | 116 |
| Pre-COVID 25 | 122 |
| Pre-COVID 26 | 120 |
| Pre-COVID 27 | 140 |
| Pre-COVID 28 | 125 |
| Pre-COVID 29 | 107 |
| Pre-COVID 30 | 130 |
| Pre-COVID 31 | 116 |
| Pre-COVID 32 | 110 |
| Pre-COVID 33 | 141 |
| Pre-COVID 34 | 128 |
| Pre-COVID 35 | 145 |
| Pre-COVID 36 | 131 |
| COVID 37 | 77 |
| COVID 38 | 121 |
| COVID 39 | 131 |
| COVID 40 | 115 |
| COVID 41 | 140 |
| COVID 42 | 141 |
| COVID 43 | 133 |
| COVID 44 | 125 |
| COVID 45 | 152 |
| COVID 46 | 192 |
| COVID 47 | 176 |
| COVID 48 | 129 |
| COVID 49 | 140 |
| COVID 50 | 176 |
| COVID 51 | 178 |
| COVID 52 | 131 |
| COVID 53 | 150 |
| COVID 54 | 160 |
| COVID 55 | 237 |
| COVID 56 | 200 |
| COVID 57 | 144 |
| COVID 58 | 180 |
| COVID 59 | 179 |
| COVID 60 | 163 |
| COVID 61 | 134 |
| COVID 62 | 178 |
| COVID 63 | 156 |
| COVID 64 | 150 |
| COVID 65 | 178 |
| COVID 66 | 150 |
| COVID 67 | 165 |
| COVID 68 | 182 |
| COVID 69 | 193 |
| COVID 70 | 187 |
| COVID 71 | 199 |
| COVID 72 | 154 |

Figure 1b: Non-COVID ICU Mortality (%)

ICU Deaths (%)

|  | ICU Death % |
| --- | --- |
| Pre-COVID 1 | 11.8782% |
| Pre-COVID 2 | 10.9091% |
| Pre-COVID 3 | 9.79878% |
| Pre-COVID 4 | 11.1217% |
| Pre-COVID 5 | 12.1557% |
| Pre-COVID 6 | 9.77948% |
| Pre-COVID 7 | 11.7867% |
| Pre-COVID 8 | 10.2452% |
| Pre-COVID 9 | 10.2383% |
| Pre-COVID 10 | 13.1533% |
| Pre-COVID 11 | 13.913% |
| Pre-COVID 12 | 11.5423% |
| Pre-COVID 13 | 12.7354% |
| Pre-COVID 14 | 10.3196% |
| Pre-COVID 15 | 11.229% |
| Pre-COVID 16 | 11.5245% |
| Pre-COVID 17 | 9.10764% |
| Pre-COVID 18 | 9.54466% |
| Pre-COVID 19 | 12.2066% |
| Pre-COVID 20 | 10.3535% |
| Pre-COVID 21 | 12.5798% |
| Pre-COVID 22 | 10.705% |
| Pre-COVID 23 | 11.1011% |
| Pre-COVID 24 | 11.5079% |
| Pre-COVID 25 | 10.7206% |
| Pre-COVID 26 | 10.7431% |
| Pre-COVID 27 | 12.2164% |
| Pre-COVID 28 | 11.5207% |
| Pre-COVID 29 | 9.70082% |
| Pre-COVID 30 | 11.8182% |
| Pre-COVID 31 | 10.1754% |
| Pre-COVID 32 | 9.42588% |
| Pre-COVID 33 | 12.3793% |
| Pre-COVID 34 | 11.2084% |
| Pre-COVID 35 | 12.0632% |
| Pre-COVID 36 | 12.5841% |
| COVID 37 | 12.4595% |
| COVID 38 | 12.8042% |
| COVID 39 | 12.3236% |
| COVID 40 | 10.6187% |
| COVID 41 | 11.9966% |
| COVID 42 | 12.6571% |
| COVID 43 | 12.3377% |
| COVID 44 | 11.2714% |
| COVID 45 | 14.0481% |
| COVID 46 | 15.534% |
| COVID 47 | 15.3444% |
| COVID 48 | 13.2853% |
| COVID 49 | 12.0069% |
| COVID 50 | 13.6646% |
| COVID 51 | 13.4238% |
| COVID 52 | 11.6652% |
| COVID 53 | 13.5379% |
| COVID 54 | 14.2857% |
| COVID 55 | 17.5816% |
| COVID 56 | 17.301% |
| COVID 57 | 13.2353% |
| COVID 58 | 15.6522% |
| COVID 59 | 15.2991% |
| COVID 60 | 15.7946% |
| COVID 61 | 12.7135% |
| COVID 62 | 15.9355% |
| COVID 63 | 13.8544% |
| COVID 64 | 13.4288% |
| COVID 65 | 16.065% |
| COVID 66 | 14.6199% |
| COVID 67 | 15.0685% |
| COVID 68 | 15.7576% |
| COVID 69 | 16.1642% |
| COVID 70 | 14.996% |
| COVID 71 | 17.1404% |
| COVID 72 | 14.4873% |

Figure 1c: Non-COVID Hospital Mortality (counts)

Hospital deaths (n)

| Month | Hospital death |
| --- | --- |
| Pre-COVID 1 | 166 |
| Pre-COVID 2 | 189 |
| Pre-COVID 3 | 161 |
| Pre-COVID 4 | 155 |
| Pre-COVID 5 | 182 |
| Pre-COVID 6 | 151 |
| Pre-COVID 7 | 167 |
| Pre-COVID 8 | 160 |
| Pre-COVID 9 | 172 |
| Pre-COVID 10 | 211 |
| Pre-COVID 11 | 209 |
| Pre-COVID 12 | 177 |
| Pre-COVID 13 | 212 |
| Pre-COVID 14 | 171 |
| Pre-COVID 15 | 178 |
| Pre-COVID 16 | 159 |
| Pre-COVID 17 | 136 |
| Pre-COVID 18 | 146 |
| Pre-COVID 19 | 162 |
| Pre-COVID 20 | 171 |
| Pre-COVID 21 | 184 |
| Pre-COVID 22 | 188 |
| Pre-COVID 23 | 185 |
| Pre-COVID 24 | 174 |
| Pre-COVID 25 | 196 |
| Pre-COVID 26 | 187 |
| Pre-COVID 27 | 188 |
| Pre-COVID 28 | 177 |
| Pre-COVID 29 | 163 |
| Pre-COVID 30 | 185 |
| Pre-COVID 31 | 160 |
| Pre-COVID 32 | 159 |
| Pre-COVID 33 | 203 |
| Pre-COVID 34 | 191 |
| Pre-COVID 35 | 199 |
| Pre-COVID 36 | 171 |
| COVID 37 | 113 |
| COVID 38 | 171 |
| COVID 39 | 186 |
| COVID 40 | 152 |
| COVID 41 | 201 |
| COVID 42 | 197 |
| COVID 43 | 193 |
| COVID 44 | 182 |
| COVID 45 | 225 |
| COVID 46 | 270 |
| COVID 47 | 227 |
| COVID 48 | 188 |
| COVID 49 | 200 |
| COVID 50 | 246 |
| COVID 51 | 261 |
| COVID 52 | 190 |
| COVID 53 | 226 |
| COVID 54 | 247 |
| COVID 55 | 387 |
| COVID 56 | 359 |
| COVID 57 | 330 |
| COVID 58 | 348 |
| COVID 59 | 375 |
| COVID 60 | 365 |
| COVID 61 | 316 |
| COVID 62 | 363 |
| COVID 63 | 321 |
| COVID 64 | 310 |
| COVID 65 | 297 |
| COVID 66 | 284 |
| COVID 67 | 319 |
| COVID 68 | 292 |
| COVID 69 | 300 |
| COVID 70 | 309 |
| COVID 71 | 292 |
| COVID 72 | 253 |

Figure 1d: Non-COVID Hospital Mortality (%)

Hospital deaths (%)

| Month | Hospital death % |
| --- | --- |
| Pre-COVID 1 | 16.9% |
| Pre-COVID 2 | 18.0861% |
| Pre-COVID 3 | 14.0857% |
| Pre-COVID 4 | 14.7338% |
| Pre-COVID 5 | 17.284% |
| Pre-COVID 6 | 14.4775% |
| Pre-COVID 7 | 15.6221% |
| Pre-COVID 8 | 14.0105% |
| Pre-COVID 9 | 15.1809% |
| Pre-COVID 10 | 18.3798% |
| Pre-COVID 11 | 18.1739% |
| Pre-COVID 12 | 17.6119% |
| Pre-COVID 13 | 19.0135% |
| Pre-COVID 14 | 15.6164% |
| Pre-COVID 15 | 15.7383% |
| Pre-COVID 16 | 14.4283% |
| Pre-COVID 17 | 12.5115% |
| Pre-COVID 18 | 12.7846% |
| Pre-COVID 19 | 15.2113% |
| Pre-COVID 20 | 14.3939% |
| Pre-COVID 21 | 16.773% |
| Pre-COVID 22 | 16.3621% |
| Pre-COVID 23 | 16.6968% |
| Pre-COVID 24 | 17.2619% |
| Pre-COVID 25 | 17.2232% |
| Pre-COVID 26 | 16.7413% |
| Pre-COVID 27 | 16.4049% |
| Pre-COVID 28 | 16.3134% |
| Pre-COVID 29 | 14.7779% |
| Pre-COVID 30 | 16.8182% |
| Pre-COVID 31 | 14.0351% |
| Pre-COVID 32 | 13.6247% |
| Pre-COVID 33 | 17.8227% |
| Pre-COVID 34 | 16.725% |
| Pre-COVID 35 | 16.5557% |
| Pre-COVID 36 | 16.4265% |
| COVID 37 | 18.2848% |
| COVID 38 | 18.0952% |
| COVID 39 | 17.4976% |
| COVID 40 | 14.0351% |
| COVID 41 | 17.2237% |
| COVID 42 | 17.684% |
| COVID 43 | 17.9035% |
| COVID 44 | 16.4112% |
| COVID 45 | 20.7948% |
| COVID 46 | 21.8447% |
| COVID 47 | 19.7908% |
| COVID 48 | 19.3615% |
| COVID 49 | 17.1527% |
| COVID 50 | 19.0994% |
| COVID 51 | 19.6833% |
| COVID 52 | 16.919% |
| COVID 53 | 20.3971% |
| COVID 54 | 22.0536% |
| COVID 55 | 28.7092% |
| COVID 56 | 31.0554% |
| COVID 57 | 30.3309% |
| COVID 58 | 30.2609% |
| COVID 59 | 32.0513% |
| COVID 60 | 35.3682% |
| COVID 61 | 29.981% |
| COVID 62 | 32.4978% |
| COVID 63 | 28.508% |
| COVID 64 | 27.7529% |
| COVID 65 | 26.8051% |
| COVID 66 | 27.6803% |
| COVID 67 | 29.1324% |
| COVID 68 | 25.2814% |
| COVID 69 | 25.1256% |
| COVID 70 | 24.7795% |
| COVID 71 | 25.1507% |
| COVID 72 | 23.8006% |

**Table 3: Subgroups and admission outcome**

| **ICU mortality** | | | | | | | | | | | | |
| --- | --- | --- | --- | --- | --- | --- | --- | --- | --- | --- | --- | --- |
|  | **Pre-pandemic period**  **n=40,196** | | | |  | **Non COVID-pandemic period**  **n=40,344** | | | |  | **For pandemic -pre-pandemic** | |
|  | **n** | **%** | **OR (95% CI)** | p value |  | **n** | **%** | **OR (95% CI)** | p value |  | **OR (95% CI)** | p value |
| **Age** |  |  |  | <0.001 |  |  |  |  | <0.001 |  |  |  |
| 18-29 | 228 | 7.7% |  |  |  | 282 | 10.1% |  |  |  | 1.15 (1.06, 1.25) | 0.001 |
| 30-39 | 312 | 9.0% |  |  |  | 465 | 12.0% |  |  |  | 1.15 (1.08, 1.23) | <0.001 |
| 40-49 | 387 | 9.0% |  |  |  | 565 | 11.8% |  |  |  | 1.14 (1.08, 1.21) | <0.001 |
| 50-59 | 775 | 10.1% |  |  |  | 950 | 12.3% |  |  |  | 1.11 (1.06, 1.16) | <0.001 |
| 60-69 | 1181 | 11.2% |  |  |  | 1574 | 14.5% |  |  |  | 1.15 (1.11, 1.19) | <0.001 |
| 70-79 | 1078 | 13.2% |  |  |  | 1413 | 17.6% |  |  |  | 1.17 (1.13, 1.22) | <0.001 |
| 80-89 | 479 | 16.9% |  |  |  | 453 | 21.4% |  |  |  | 1.17 (1.09, 1.26) | <0.001 |
| 90+ | 63 | 24.9% |  |  |  | 25 | 17.4% |  |  |  | 0.74 (0.51, 1.06) | 0.082 |
| **Sex** |  |  | 1.07 (1.03, 1.11) | <0.001 |  |  |  | 1.04 (1.00, 1.08) | 0.026 |  |  |  |
| Female | 1851 | 11.9% |  |  |  | 2231 | 14.7% |  |  |  | 1.12 (1.09, 1.16) | <0.001 |
| Male | 2652 | 10.8% |  |  |  | 3494 | 13.9% |  |  |  | 1.15 (1.12, 1.17) | <0.001 |
| **APACHE II score** |  |  | 1.88 (1.85, 1.90) | <0.001 |  |  |  | 1.73 (1.70, 1.75) | <0.001 |  |  |  |
| APACHE II score < 17 | 208 | 1.2% |  |  |  | 476 | 2.9% |  |  |  | 1.45 (1.37, 1.52) | <0.001 |
| APACHE II score ≥ 17 | 3755 | 17.4% |  |  |  | 4796 | 21.0% |  |  |  | 1.11 (1.09, 1.14) | <0.001 |
| **APACHE III score** |  |  | 1.86 (1.83, 1.88) | <0.001 |  |  |  | 1.82 (1.79, 1.84) | <0.001 |  |  |  |
| APACHE III score < 55 | 231 | 1.3% |  |  |  | 479 | 2.7% |  |  |  | 1.36 (1.29, 1.44) | <0.001 |
| APACHE III score ≥ 55 | 4265 | 19.0% |  |  |  | 5184 | 23.0% |  |  |  | 1.12 (1.10, 1.15) | <0.001 |
| **SOFA score** |  |  | 1.49 (1.47, 1.50) | <0.001 |  |  |  | 1.45 (1.43, 1.46) | <0.001 |  |  |  |
| SOFA score < 5 | 278 | 2.1% |  |  |  | 593 | 4.3% |  |  |  | 1.36 (1.30, 1.43) | <0.001 |
| SOFA score ≥ 5 | 4223 | 15.8% |  |  |  | 5083 | 19.2% |  |  |  | 1.12 (1.10, 1.14) | <0.001 |
| **CFS score** |  |  | 1.80 (1.73, 1.87) | <0.001 |  |  |  | 1.82 (1.74, 1.90) | <0.001 |  |  |  |
| CFS score < 5 | 2255 | 9.0% |  |  |  | 2817 | 11.7% |  |  |  | 1.15 (1.12, 1.18) | <0.001 |
| CFS score ≥ 5 | 1730 | 19.2% |  |  |  | 1736 | 23.4% |  |  |  | 1.14 (1.10, 1.19) | <0.001 |
| **Hospital mortality** | | | | | | | | | | | | |
|  | **Pre-pandemic period**  **n=40,196** | | | |  | **Non COVID-pandemic period**  **n=40,344** | | | |  | **For pre-pandemic -pandemic** | |
|  | **n** | **%** | **OR (95% CI)** | **p value** |  | **n** | **%** | **OR (95% CI)** | **p value** |  | **OR (95% CI)** | **p value** |
| **Age** |  |  |  | <0.001 |  |  |  |  | <0.001 |  |  |  |
| 18-29 | 278 | 9.4% |  |  |  | 433 | 15.5% |  |  |  | 1.30 (1.22, 1.39) | <0.001 |
| 30-39 | 383 | 11.0% |  |  |  | 693 | 17.8% |  |  |  | 1.27 (1.21, 1.33) | <0.001 |
| 40-49 | 521 | 12.1% |  |  |  | 939 | 19.6% |  |  |  | 1.28 (1.22, 1.33) | <0.001 |
| 50-59 | 1060 | 13.8% |  |  |  | 1664 | 21.6% |  |  |  | 1.28 (1.23, 1.32) | <0.001 |
| 60-69 | 1732 | 16.4% |  |  |  | 2627 | 24.1% |  |  |  | 1.25 (1.21, 1.28) | <0.001 |
| 70-79 | 1607 | 19.7% |  |  |  | 2347 | 29.2% |  |  |  | 1.28 (1.24, 1.32) | <0.001 |
| 80-89 | 734 | 26.0% |  |  |  | 790 | 37.3% |  |  |  | 1.34 (1.25, 1.42) | <0.001 |
| 90+ | 87 | 34.4% |  |  |  | 44 | 30.6% |  |  |  | 0.89 (0.67, 1.19) | 0.435 |
| **Sex** |  |  | 1.04 (1.0, 1.08) | 0.016 |  |  |  | 1.05 (1.02, 1.08) | 0.002 |  |  |  |
| Female | 2560 | 16.5% |  |  |  | 3718 | 24.5% |  |  |  | 1.26 (1.23, 1.29) | <0.001 |
| Male | 3840 | 15.6% |  |  |  | 5817 | 23.1% |  |  |  | 1.25 (1.23, 1.28) | <0.001 |
| **APACHE II score** |  |  | 1.86 (1.83, 1.89) | <0.001 |  |  |  | 1.71 (1.68, 1.73) | <0.001 |  |  |  |
| APACHE II score < 17 | 547 | 3.1% |  |  |  | 1361 | 8.2% |  |  |  | 1.51 (1.46, 1.56) | <0.001 |
| APACHE II score ≥ 17 | 5272 | 24.5% |  |  |  | 7671 | 33.6% |  |  |  | 1.23 (1.21, 1.25) | <0.001 |
| **APACHE III score** |  |  | 1.85 (1.82, 1.87) | <0.001 |  |  |  | 1.79 (1.77, 1.82) | <0.001 |  |  |  |
| APACHE III score < 55 | 561 | 3.2% |  |  |  | 1432 | 8.1% |  |  |  | 1.48 (1.44, 1.53) | <0.001 |
| APACHE III score ≥ 55 | 5832 | 25.9% |  |  |  | 8029 | 35.6% |  |  |  | 1.24 (1.22, 1.27) | <0.001 |
| **SOFA score** |  |  | 1.43 (1.41, 1.44) | <0.001 |  |  |  | 1.43 (1.41, 1.45) | <0.001 |  |  |  |
| SOFA score < 5 | 708 | 5.3% |  |  |  | 1384 | 10.1% |  |  |  | 1.34 (1.30, 1.39) | <0.001 |
| SOFA score ≥ 5 | 5691 | 21.3% |  |  |  | 8089 | 30.5% |  |  |  | 1.26 (1.24, 1.28) | <0.001 |
| **CFS score** |  |  | 2.02 (1.95, 2.10) | <0.001 |  |  |  | 2.00 (1.93, 2.08) | <0.001 |  |  |  |
| CFS score < 5 | 3065 | 12.2% |  |  |  | 4799 | 19.9% |  |  |  | 1.31 (1.28, 1.33) | <0.001 |
| CFS score ≥ 5 | 2580 | 28.6% |  |  |  | 2908 | 39.3% |  |  |  | 1.29 (1.25, 1.33) | <0.001 |

* p value <0.001; ‡ p value=0.01; $ p value=0.002
APACHE: Acute Physiologic Assessment and Chronic Health Evaluation CFS: Clinical Frailty Scale, CI: confidence intervals, ICU: intensive care unit, OR: odds ratio, SD: standard deviation, SOFA: Sequential Organ Failure Assessment

**Figure 2: Subgroup analysis**

Figure 2a: ICU Mortality

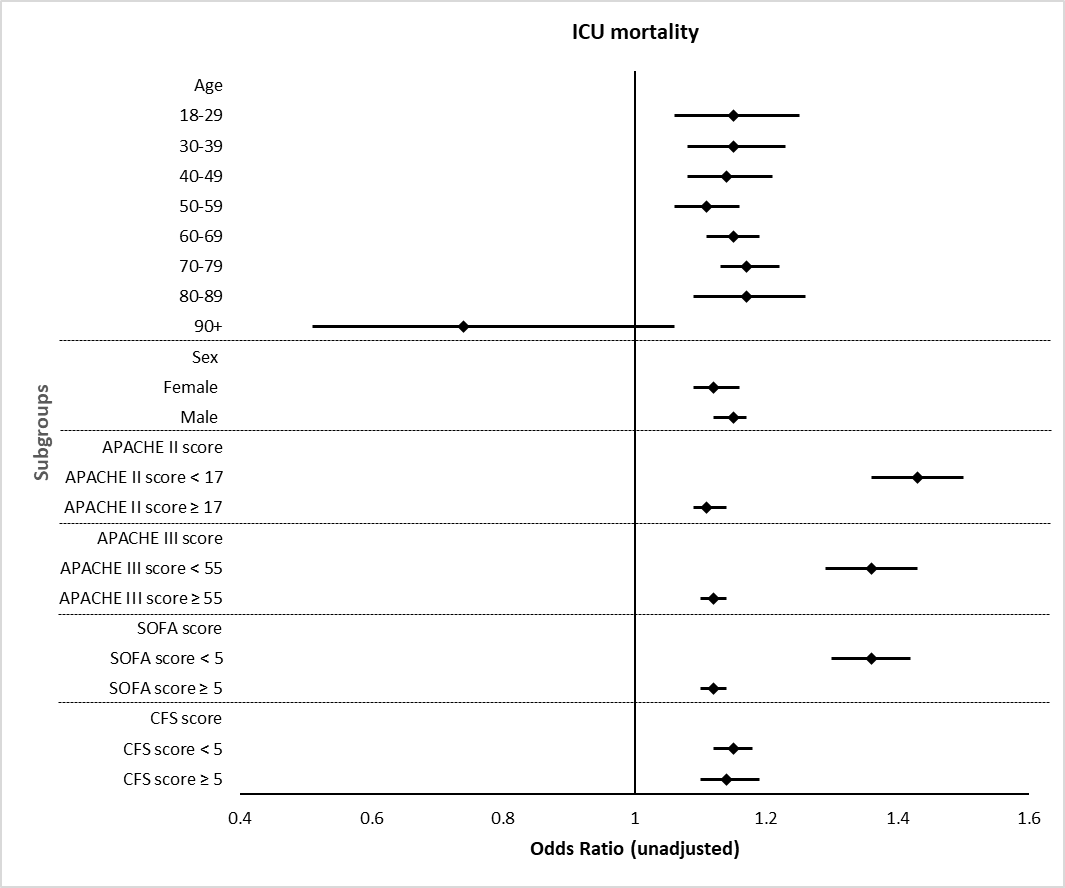

Figure 2b: Hospital Mortality

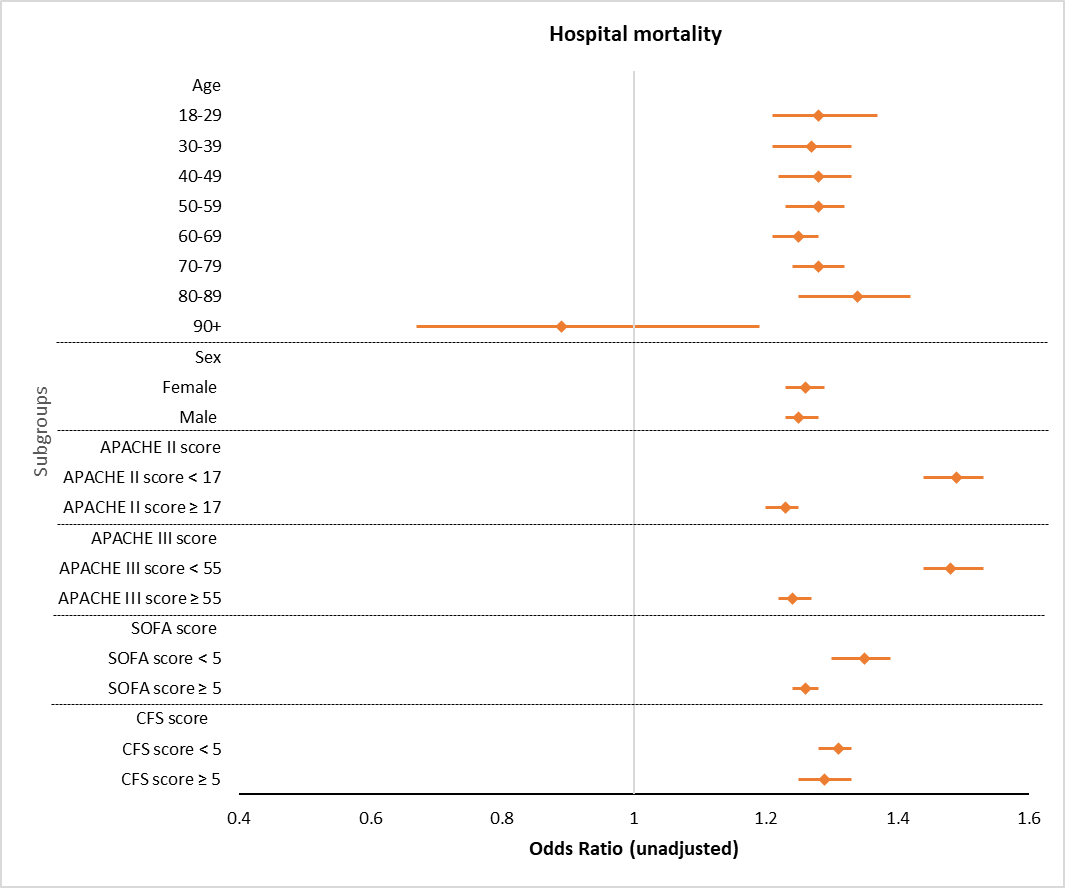

**Supplementary materials**

**Supplemental Table 1: Patients’ characteristics in propensity-matched cohort**

|  |  | **Pre-pandemic period**  **n=15,614** | | | | |  | **Non COVID-pandemic period**  **n=15,614** | | | | |  | **p value** |
| --- | --- | --- | --- | --- | --- | --- | --- | --- | --- | --- | --- | --- | --- | --- |
| **Sex** |  |  | | |  | |  |  | | |  | |  | 0.17 |
| Female |  | 5377 | | | 34.4% | |  | 5493 | | | 35.2% | |  |  |
| Male |  | 10237 | | | 65.6% | |  | 10121 | | | 64.8% | |  |  |
| **Age** |  |  | | |  | |  |  | | |  | |  |  |
| 18-29 |  | 1373 | | | 8.8% | |  | 1331 | | | 8.5% | |  | 0.42 |
| 30-39 |  | 1749 | | | 11.2% | |  | 1720 | | | 11.0% | |  | 0.62 |
| 40-49 |  | 2104 | | | 13.5% | |  | 1963 | | | 12.6% | |  | 0.03 |
| 50-59 |  | 3440 | | | 22.0% | |  | 3277 | | | 21.0% | |  | 0.05 |
| 60-69 |  | 4165 | | | 26.7% | |  | 4154 | | | 26.6% | |  | 0.90 |
| 70-79 |  | 2357 | | | 15.1% | |  | 2690 | | | 17.2% | |  | **<0.001** |
| 80-89 |  | 417 | | | 2.7% | |  | 473 | | | 3.0% | |  | 0.06 |
| 90+ |  | 9 | | | 0.1% | |  | 6 | | | 0.0% | |  | 0.44 |
| **Comorbidities** |  |  | | |  | |  |  | | |  | |  |  |
| Dialysis |  | 339 | | | 2.2% | |  | 305 | | | 2.0% | |  | 0.18 |
| Heart Disease |  | 1364 | | | 8.7% | |  | 1392 | | | 8.9% | |  | 0.58 |
| Respiratory insufficiency |  | 1238 | | | 7.9% | |  | 1255 | | | 8.0% | |  | 0.72 |
| Liver disease |  | 888 | | | 5.7% | |  | 894 | | | 5.7% | |  | 0.88 |
| Cirrhosis |  | 789 | | | 5.1% | |  | 782 | | | 5.0% | |  | 0.87 |
| Hepatic failure |  | 425 | | | 2.7% | |  | 428 | | | 2.7% | |  | 0.91 |
| Diabetes |  | 3399 | | | 21.8% | |  | 3345 | | | 21.4% | |  | 0.46 |
| Leukemia/M. Myeloma |  | 93 | | | 0.6% | |  | 66 | | | 0.4% | |  | **0.04** |
| Lymphoma |  | 91 | | | 0.06 | |  | 73 | | | 0.5% | |  | 0.18 |
| Immune suppression |  | 1008 | | | 6.5% | |  | 983 | | | 6.3% | |  | 0.57 |
| Metastatic cancer |  | 332 | | | 2.1% | |  | 289 | | | 1.9% | |  | 0.09 |
| **Admission class** |  |  | | |  | |  |  | | |  | |  |  |
| Medical |  | 6153 | | | 39.4% | |  | 6131 | | | 39.3% | |  | 0.84 |
| Surgical |  | 7412 | | | 47.5% | |  | 7495 | | | 48.0% | |  | 0.50 |
| Neuro |  | 1106 | | | 7.1% | |  | 1093 | | | 7.0% | |  | 0.78 |
| Trauma and head injury |  | 476 | | | 3.0% | |  | 463 | | | 3.0% | |  | 0.67 |
| Trauma without head injury |  | 467 | | | 3.0% | |  | 432 | | | 2.8% | |  | 0.24 |
| **APACHE II score** |  |  | | |  | |  |  | | |  | |  | 0.98 |
| < 17 |  | 6257 | | | 40.1% | |  | 6259 | | | 40.1% | |  |  |
| ≥ 17 |  | 9357 | | | 59.9% | |  | 9355 | | | 59.9% | |  |  |
|  |  | **Pre-pandemic period**  **n=15,614** | | | | |  | **Non COVID-pandemic period**  **n=15,614** | | | | |  | **p value**† |
|  |  | **n** | **Mean** | **(SD)** | | **Median [IQR]** |  | **n** | **Mean** | **(SD)** | | **Median [IQR]** |  |  |
| **Age (year)** |  | 15614 | 54.70 | (15.85) | | 57 [44, 67] |  | 15614 | 55.46 | (15.98) | | 58 [44, 68] |  | **<0.001** |
| **Weight (KG)** |  | 15168 | 84.46 | (24.53) | | 81 [69.1, 97] |  | 10354 | 85.64 | (24.86) | | 82 [69, 97.9] |  | 0.24 |
| **APACHE-II** |  | 15614 | 20.02 | (8.54) | | 19 [14, 25] |  | 15614 | 20.04 | (8.44) | | 18 [14, 25] |  | 0.57 |
| **Invasive ventilation (day)** |  | 15614 | 3.13 | (4.60) | | 1.09 [0.39, 3.54] |  | 15614 | 2.97 | (4.63) | | 1.25 [0.44, 3.84] |  | **<0.001** |

APACHE: Acute Physiologic Assessment and Chronic Health Evaluation, CFS: Clinical Frailty Scale, CI: confidence intervals, COVID: Coronavirus Disease, ICU: intensive care unit, KG: kilograms, SD: standard deviation, SOFA: Sequential Organ Failure Assessment, † Mann-Whitney U test

**Supplemental Table 2: Patients’ characteristics in propensity-unmatched cohort**

|  |  | **Pre-pandemic period**  **n=24,582** | | | | |  | | **Non COVID-pandemic period**  **n=24,730** | | | | | |  | **p value** |
| --- | --- | --- | --- | --- | --- | --- | --- | --- | --- | --- | --- | --- | --- | --- | --- | --- |
| **Sex** |  |  | | |  | |  | |  | | | |  | |  | **<0.001** |
| Female |  | 10160 | | | 41.3% | |  | | 9691 | | | | 39.2% | |  |  |
| Male |  | 14416 | | | 58.6% | |  | | 15034 | | | | 60.8% | |  |  |
| Unknown |  | 6 | | | 0.0% | |  | | 5 | | | | 0.0% | |  |  |
| **Age** |  |  | | |  | |  | |  | | | |  | |  | **<0.001** |
| 18-29 |  | 1579 | | | 6.4% | |  | | 1457 | | | | 5.9% | |  | **0.02** |
| 30-39 |  | 1729 | | | 7.0% | |  | | 2168 | | | | 8.8% | |  | **<0.001** |
| 40-49 |  | 2203 | | | 9.0% | |  | | 2826 | | | | 11.4% | |  | **<0.001** |
| 50-59 |  | 4234 | | | 17.2% | |  | | 4437 | | | | 17.9% | |  | 0.06 |
| 60-69 |  | 6386 | | | 26.0% | |  | | 6724 | | | | 27.2% | |  | **0.009** |
| 70-79 |  | 5796 | | | 23.6% | |  | | 5335 | | | | 21.6% | |  | **<0.001** |
| 80-89 |  | 2411 | | | 9.8% | |  | | 1645 | | | | 6.7% | |  | **<0.001** |
| 90+ |  | 244 | | | 1.0% | |  | | 138 | | | | 0.6% | |  | **<0.001** |
| **Comorbidities** |  |  | | |  | |  | |  | | | |  | |  |  |
| Dialysis |  | 771 | | | 3.5% | |  | | 643 | | | | 3.6% | |  | 0.53 |
| Heart Disease |  | 2238 | | | 10.1% | |  | | 1798 | | | | 10.1% | |  | 0.88 |
| Respiratory insufficiency |  | 2998 | | | 13.6% | |  | | 2192 | | | | 12.3% | |  | **<0.001** |
| Liver disease |  | 1047 | | | 4.7% | |  | | 1123 | | | | 6.3% | |  | **<0.001** |
| Cirrhosis |  | 913 | | | 4.1% | |  | | 983 | | | | 5.5% | |  | **<0.001** |
| Hepatic failure |  | 484 | | | 2.2% | |  | | 522 | | | | 2.9% | |  | **<0.001** |
| Diabetes |  | 5494 | | | 24.9% | |  | | 4836 | | | | 27.2% | |  | **<0.001** |
| Leukemia/M. Myeloma |  | 209 | | | 0.9% | |  | | 177 | | | | 1.0% | |  | 0.63 |
| Lymphoma |  | 221 | | | 1.0% | |  | | 183 | | | | 1.0% | |  | 0.80 |
| Immune suppression |  | 1810 | | | 8.2% | |  | | 1624 | | | | 9.1% | |  | **0.001** |
| Metastatic cancer |  | 772 | | | 3.5% | |  | | 556 | | | | 3.1% | |  | **0.04** |
| **Admission class** |  |  | | |  | |  | |  | | | |  | |  | **<0.001** |
| Medical |  | 11987 | | | 54.3% | |  | | 12406 | | | | 69.8% | |  | **<0.001** |
| Surgical |  | 7807 | | | 35.4% | |  | | 3901 | | | | 21.9% | |  | **<0.001** |
| Neuro |  | 1607 | | | 7.3% | |  | | 1004 | | | | 5.6% | |  | **<0.001** |
| Trauma and head injury |  | 408 | | | 1.8% | |  | | 250 | | | | 1.4% | |  | **<0.001** |
| Trauma without head injury |  | 258 | | | 1.2% | |  | | 217 | | | | 1.2% | |  | 0.6 |
| **APACHE II score** |  |  | | |  | |  | |  | | | |  | |  | **<0.001** |
| < 17 |  | 11396 | | | 48.4% | |  | | 10419 | | | | 43.6% | |  |  |
| ≥ 17 |  | 12173 | | | 51.6% | |  | | 13478 | | | | 56.4% | |  |  |
|  |  | **Pre-pandemic period**  **n=24,582** | | | | | |  | | **Non COVID-pandemic period**  **n=24,730** | | | | |  | **p value**† |
|  |  | **n** | **Mean** | **(SD)** | | **Median [IQR]** | |  | | **n** | **Mean** | **(SD)** | | **Median [IQR]** |  |  |
| **Age (year)** |  | 24582 | 60.89 | (16.61) | | 64 [52, 73] | |  | | 24730 | 59.03 | (16.12) | | 62 [49, 73] |  | **<0.001** |
| **Weight (KG)** |  | 23382 | 84.15 | (25.06) | | 80 [67.6, 96] | |  | | 13747 | 86.53 | (26.28) | | 82.5 [69, 99] |  | **<0.001** |
| **APACHE-II** |  | 23569 | 18.05 | (8.29) | | 17 [12, 23] | |  | | 23897 | 19.15 | (9.04) | | 18 [12, 25] |  | **<0.001** |
| **Invasive ventilation (day)** |  | 11225 | 3.85 | (9.31) | | 0.87 [0.29, 3.39] | |  | | 13307 | 6.04 | (10.43) | | 2.09 [0.61, 7.07] |  | **<0.001** |

**Supp. Table 3: Distribution of ICU admissions by hospital ICU**

| **Site** | **Pre-pandemic period**  **n=40,196** | | **Non COVID-pandemic period**  **n=40,344** | |
| --- | --- | --- | --- | --- |
| Alberta Children’s Pediatric ICU (Calgary) | 11 | 0.0% | **17** | **0.0%** |
| Chinook Regional Hospital ICU (Calgary) | **1924** | **4.8%** | 1579 | 3.9% |
| Foothills Medical Center Cardiovascular ICU (Calgary) | **4607** | **11.5%** | 4383 | 10.9% |
| Foothill Medical Center ICU (Calgary) | 3787 | 9.4% | **4438** | **11.0%** |
| Grey Nun’s Hospital ICU (Edmonton) | 1372 | 3.4% | **1511** | **3.7%** |
| Mazankowski Heart Institute Cardiovascular ICU (Edmonton) | **4828** | **12.0%** | 4374 | 10.8% |
| Medicine Hat Regional Hospital ICU (Medicine Hat) | **1300** | **3.2%** | 972 | 2.4% |
| Misericordia Hospital ICU (Edmonton) | **1266** | **3.1%** | 1192 | 3.0% |
| Northern Lights Health Center ICU (Fort McMurray) | **1089** | **2.7%** | 887 | 2.2% |
| Peter Lougheed Center ICU (Calgary) | 2105 | 5.2% | **2539** | **6.3%** |
| Queen Elizabeth II ICU (Grande Prairie) | 1108 | 2.8% | **1179** | **2.9%** |
| Royal Alexandra Hospital ICU (Edmonton) | 4228 | 10.5% | **4353** | **10.8%** |
| Red Deer Regional Hospital ICU (Red Deer) | 1732 | 4.3% | **1788** | **4.4%** |
| Rockyview Hospital ICU (Calgary) | 1441 | 3.6% | 1453 | 3.6% |
| Sturgeon Community Hospital ICU (St. Albert) | 999 | 2.5% | **1060** | **2.6%** |
| South Health Campus ICU (Calgary) | 1241 | 3.1% | **1482** | **3.7%** |
| Stollery Pediatric ICU (Edmonton) | 0 | 0.0% | **14** | **0.0%** |
| University of Alberta Hospital Burns ICU (Edmonton) | 64 | 0.2% | **332** | **0.8%** |
| University of Alberta Hospital General Systems ICU (Edmonton) | 4948 | 12.3% | 4991 | 12.4% |
| University of Alberta Hospital Neurological ICU (Edmonton) | **2158** | **5.4%** | 1820 | 4.5% |

ICU: intensive care unit

**Supplemental Table 4: Admission reason**

|  | **Pre-pandemic period**  **n=40,196** | | **Non COVID-pandemic period**  **n=40,344** | |
| --- | --- | --- | --- | --- |
| **Cardiovascular Non-Operative** | 64 | 0.2% | **900** | **2.2%** |
| **Cardiovascular Post-Operative** | 391 | 1.0% | **3484** | **8.6%** |
| **Elective surgery** | **9232** | **23.0%** | 3483 | 8.6% |
| **Emergency** | **5662** | **14.1%** | 3224 | 8.0% |
| **Gastrointestinal (GI) Non-Operative** | 45 | 0.1% | **583** | **1.4%** |
| **Gastrointestinal (GI) Post-Operative** | 32 | 0.1% | **369** | **0.9%** |
| **Genitourinary Non-Operative** | 8 | 0.0% | **171** | **0.4%** |
| **Genitourinary Post-Operative** | 11 | 0.0% | **70** | **0.2%** |
| **Hematology Non-Operative** | 2 | 0.0% | **31** | **0.1%** |
| **Hematology Post-Operative** | 0 | 0.0% | 1 | 0.0% |
| **Metabolic/Endocrine Non-Operative** | 7 | 0.0% | **149** | **0.4%** |
| **Metabolic/Endocrine Post-Operative** | 0 | 0.0% | **7** | 0.0% |
| **Musculoskeletal/Skin Non-Operative** | 10 | 0.0% | **121** | **0.3%** |
| **Musculoskeletal/Skin Post-Operative** | 10 | 0.0% | **164** | **0.4%** |
| **Neurologic Non-Operative** | 63 | 0.2% | **1116** | **2.8%** |
| **Neurologic Post-Operative** | 31 | 0.1% | **337** | **0.8%** |
| **Non-Operative** | **21867** | **54.4%** | 15865 | 39.3% |
| **Orthopedic surgery** | **2** | **0.0%** | 1 | 0.0% |
| **Respiratory Non-Operative** | 108 | 0.3% | **1938** | **4.8%** |
| **Respiratory Post-Operative** | 43 | 0.1% | **186** | **0.5%** |
| **Transplant Non-Operative** | 0 | 0.0% | 5 | 0.0% |
| **Transplant Post-Operative** | 44 | 0.1% | **365** | **0.9%** |
| **Trauma Non-Operative** | 23 | 0.1% | **380** | **0.9%** |
| **Trauma Post-Operative** | 5 | 0.0% | **253** | **0.6%** |
| **Missing** | 2536 | 6.3% | **7141** | **17.7%** |

**Supplemental Table 5: Resource utilization and organ supports**

|  |  | **Pre-pandemic period**  **n=40,196** | | | |  | **Non COVID-pandemic period**  **n=40,344** | | | |  |  | |
| --- | --- | --- | --- | --- | --- | --- | --- | --- | --- | --- | --- | --- | --- |
|  |  | **n** | **(%)** |  |  |  | **n** | **(%)** |  |  |  | ***X2*** | **p value** |
| **Airway pressure release ventilation** |  | 168 | 0.4% | **--** | **--** |  | 423 | 1.0% | **--** | **--** |  | 109.90 | <0.001 |
| **Sustained low-efficiency dialysis** |  | 79 | 4.7% | **--** | **--** |  | 22 | 0.1% | **--** | **--** |  | 562.26 | <0.001 |
| **Tracheostomy** |  | 2570 | 6.4% | **--** | **--** |  | 2815 | 7.0% | **--** | **--** |  | 11.00 | <0.001 |
| **Extracorporeal membrane oxygenation** |  | 216 | 0.5% | **--** | **--** |  | 339 | 0.8% | **--** | **--** |  | 27.00 | <0.001 |
| **Unplanned Extubating** |  | 407 | 1.0% | **--** | **--** |  | 270 | 0.7% | **--** | **--** |  | 28.47 | <0.001 |
|  |  | **n** | **(%)** | **Median** | **[IQR]** |  | **n** | **(%)** | **Median** | **[IQR]** |  | **p value**† | |
| **Invasive ventilation (day)** |  | 26839 | 66.8% | 1.00 | [0.3, 3.5] |  | 28921 | 71.7% | **1.6** | [0.5, 5.1] |  | <0.001 | |
| **Non-Invasive ventilation (day)** |  | 4387 | 10.9% | 0.58 | [0.2, 1.3] |  | 2780 | 6.9% | 0.66 | [0.2, 1.4] |  | 0.001 | |
| **Continuous renal replacement therapy (day)** |  | 2016 | 5.0% | 2.8 | [1.2, 5.8] |  | 2019 | 5.0% | 3.0 | [1.4, 6.4] |  | 0.001 | |
| **Intermittent hemodialysis (hours)** |  | 1634 | 4.1% | 8.4 | [4.3, 17.2] |  | 924 | 2.3% | 8.2 | [4.2, 17.1] |  | 0.17 | |
| **Vasopressors used** |  |  |  |  |  |  |  |  |  |  |  |  | |
| Dopamine (hours) |  | 2399 | 6.0% | 2.65 | [0.77, 11.3] |  | 1663 | 4.1% | **3.50** | [0.87, 15.25] |  | <0.001 | |
| Dobutamine (hours) |  | 2218 | 5.5% | 29.17 | [7.85, 63.28] |  | 2199 | 5.5% | **33.35** | [11.98, 69.00] |  | <0.001 | |
| Epinephrine (hours) |  | 2863 | 7.1% | 7.97 | [1.83, 22.47] |  | 3287 | 8.1% | **8.40** | [2.18, 25.47] |  | 0.015 | |
| Norepinephrine (hours) |  | 20761 | 51.6% | 18.90 | [6.25, 44.65] |  | 24899 | 61.7% | **25.22** | [8.45, 60.05] |  | <0.001 | |
| Vasopressin (hours) |  | 6530 | 16.2% | 26.46 | [11.30, 52.28] |  | 8171 | 20.3% | 27.08 | [11.60, 53.32] |  | 0.314 | |
| Isoproterenol (hours) |  | 127 | 0.3% | **22.15** | [5.08, 48.72] |  | 147 | 0.4% | 9.97 | [1.63, 24.00] |  | <0.001 | |
| Phenylephrine (hours) |  | 4619 | 11.5% | **0.07** | [0.3, 0.17] |  | 4506 | 11.2% | 0.05 | [0.02, 0.13] |  | <0.001 | |
| Milrinone (hours) |  | 2466 | 6.1% | **39.33** | [17.70, 78.82] |  | 2249 | 5.6% | 34.87 | [15.55, 69.04] |  | <0.001 | |
| Epoprostenol (hours) |  | 926 | 2.3% | 26.96 | [11.22, 64.68] |  | 128 | 3.0% | **38.58** | [14.25, 76.97] |  | <0.001 | |

† Wilcoxon Rank-Sum test
